# Mechanistic modelling of coronavirus infections and the impact of confined neighbourhoods on a short time scale

**DOI:** 10.1101/2020.07.28.20163634

**Authors:** Danish A Ahmed, Ali R Ansari, Mudassar Imran, Kamaludin Dingle, Naveed Ahmed, Michael B Bonsall

## Abstract

**Background:** To mitigate the spread of the COVID-19 coronavirus, some countries have adopted more stringent non-pharmaceutical interventions in contrast to those widely used (for e.g. the state of Kuwait). In addition to standard practices such as enforcing curfews, social distancing, and closure of non-essential service industries, other non-conventional policies such as the total confinement of highly populated areas has also been implemented.

**Methods:** In this paper, we model the movement of a host population using a mechanistic approach based on random walks, which are either diffusive or super-diffusive. Infections are realised through a contact process, whereby a susceptible host may be infected if in close spatial proximity of the infectious host. Our focus is only on the short-time scale prior to the infectious period, so that no further transmission is assumed.

**Results:** We find that the level of infection depends heavily on the population dynamics, and increases in the case of slow population diffusion, but remains stable for a high or super-diffusive population. Also, we find that the confinement of homogeneous or overcrowded sub-populations has minimal impact in the short term.

**Conclusions:** Our results indicate that on a short time scale, confinement restrictions or complete lock down of whole residential areas may not be effective. Finally, we discuss the possible implications of our findings for total confinement in the context of the current situation in Kuwait.

## Introduction

The novel coronavirus SARS-CoV2 referred to by the World Health Organization (WHO) as COVID-19 (Coronavirus Disease 2019) is believed to have started from an animal source in Wuhan City, Hubei Province, China in December 2019 [1, 2]. Since then, the disease has spread worldwide, across 188 countries, territories and regions, making it a global health emergency [3, 4]. On 11^th^ March, the WHO officially class-fied the COVID-19 outbreak as a pandemic. As of July 22^nd^ 2020, COVID-19 has infected 14,960,136 individuals of which 616,769 deaths have occurred and 8,480,064 have recovered, making the total active infected cases 6,480,072 [3]. Whilst most symptomatic cases are mild, characterised by a persistent cough and fever, a significant proportion of cases are more serious, where individuals develop pneumonia - leading to acute respiratory failure, which can possibly be fatal. A combination of vastly different non-pharmacological measures have been adopted by national governments to suppress the growth of the epidemic, such as: travel bans, school closures, social distancing, imposed curfews, household quarantine, complete lock-downs, etc [5]. The extent to which of these strategies are most effective, including their timing, is not entirely clear. Although studies have attempted to identify key intervention policies [6, 7], others have highlighted those which are ineffective [8, 9]. COVID-19 is highly contagious and more infectious than initially thought, where improved estimates have shown that during the early stages of the epidemic spread, the number of infected individuals can double every 2.4 days [10]. The virus continues to spread in a similar way to influenza, via respiratory droplets from coughing or sneezing. Therefore, the primary mode of transmission is attained through a ‘contact’ process, i.e. if susceptible individuals are in close spatial proximity of infectious hosts [11]. Once a person is infected, for most people (approx. 81%), no symptoms will show [12]. For others, the time between exposure to the virus (becoming infected) and symptom onset, is on average 5-6 days, but can range from 2-14 days [13, 14]. It is also estimated that a virus carrier will typically only become infectious around 1-3 days before symptoms appear [15]. How long it takes, and to what extent asymptomatic individuals transmit the disease is not completely understood [16, 17]. The preceding suggests that given contact between susceptible and infectious hosts, for an initial period of many days, the only, or at least primary means of the virus spreading will be from the initial infected individual to others, without further transmission. This early period will have qualitatively different virus spreading characteristics as compared to the later stages, where newly infected individuals can also spread the disease. Hence it is of interest to study the short-time dynamics of infection levels and of the subtle interplay between the processes involved on this time scale [18, 19].

Mechanistic movement models provide an alternative modelling approach to conventional epidemiological models (SIR, SEIR), as a means to better understand the dynamics of disease spread [20, 21, 22, 11]. One advantage is that the spatial proximity between individuals is explicitly accounted for through individual movement rules, where susceptible individuals come into close contact with infectious hosts, and are possibly infected. Therefore, movement behaviours and the contact patterns that emerge due to these encounters directly relate to disease transmission. In terms of mathematical modelling, Random Walks (RWs) serve as a useful modelling tool to track the movement of individuals in a population across space and time [23, 24]. A basic description is given by the Correlated Random Walk (CRW), where the orientations between successive steps are correlated, resulting in a localized directional persistence [25, 26, 27, 28]. This means that individuals in the short term are more likely to keep moving in the same direction than to perform abrupt turns. In the absence of directional persistence, the CRW reduces to the Simple Random Walk (SRW), which can be considered as a special case, so that the movement is uncorrelated and completely random [29, 30]. In the case of a population of non-interacting individuals, such movement processes are known to be diffusive, particularly at large spatial scales [31, 32]. In movement ecology, the CRW is supported by empirical evidence from animal movement data, and thus frequently used to model animal movement paths [33, 28, 34, 35]. However, in the case of more complicated movement types, such as that observed for humans, the CRW does not provide an adequate description, but can still serve as a null model. To the best of our knowledge, no epidemiological studies have considered host movement as a CRW - even in disease ecology.

Another conceptual tool for modelling movement is the Lévy Walk (LW), where the individual performs short steps forming clusters, with the occasional longer step in between them [36, 37, 38]. If the LW is oriented during the clustering phases, the corresponding movement type is referred to as the Correlated Lévy Walk (CLW). In contrast to the CRW, the movement pattern is much faster, and super-diffusive. It is now generally accepted that some animal species perform LWs [39, 40, 41], particularly in context-specific scenarios such as foraging, and known to describe an efficient searching strategy where resources are scarce and randomly distributed [42, 43, 44]. Alongside this, there is growing empirical evidence that human movements may also exhibit Lévy type characteristics. Such inferences have been reached from studies on the daily movement patterns of humans, traces of bank notes, mobile phone users’ locations and GPS trajectories [45, 46, 47, 48, 49]. Therefore, a LW description could be useful to study a wide variety of challenging issues; such as traffic prediction, urban planning, and in the context of our study, epidemic spread [50]. Despite the clear motivation, few studies have focused on epidemics in populations where the host population performs a LW. As an example, it was demonstrated in [51] that a disease outbreak is more likely for similar density populations where individuals perform the LW, instead of the SRW.

In this paper we use a mechanistic description based on RWs to model the movement of susceptible and infectious hosts in 2D space. We consider the early stage of epidemic development, during the incubation phase prior to the infectious period, where it is assumed that the virus cannot be further transmitted. We demonstrate how different modes of host movements can lead to varying levels of infections. In addition, we consider various confinement scenarios for both homogeneous and overcrowded populations, where the movement is restricted to a certain area. Thus, we reveal whether confinement is effective in mitigating disease spread, at least on a short time scale.

## Methods

Random walk framework

The movement of a walker in 2D space along a curvilinear path in continuous space-time, **x** = **x**(*t*) = (*x*(*t*), *y*(*t*)) can be modelled using a discrete time random walk (RW) which links individual location **x**_*i−*1_ at time *t*_*i−*1_ to the next location **x**_*i*_ at time *t*_*i*_. Each location is recorded at *t*_*i*_ = *i*Δ*t* = *{t*_0_, *t*_1_, *t*_2_, …*}* where Δ*t* is considered as a constant time step, independent of *i*. The step length defined as the distance between any two successive steps is *l*_*i*_ = |**x**_*i*_ − **x**_*i−*1_| = *{l*_1_, *l*_2_, *l*_3_, …*}* with average velocity 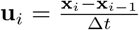 [28, 52]. The complete movement path which begins at location **x**_0_ can then be expressed through the equation:

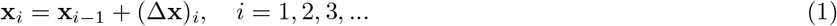

where (Δ**x**)_*i*_ = (Δ*x*_*i*_, Δ*y*_*i*_) is a random step vector for the *i*^*th*^ step along the walk. Any 2D RW can also be described in polar co-ordinates, by expressing the step vector in terms of step lengths *l* and turning angle *θ* (i.e. the angle between two consecutive headings), using the transformation:

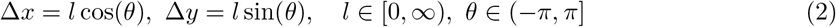

with inverse transformation:

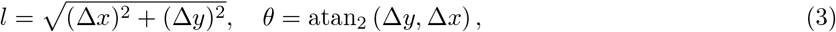

where atan_2_ (Δ*y*, Δ*x*) is equal to arctan 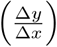 for Δ*x >* 0 and to arctan 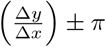 for Δ*x* < 0. The 2D RW can then be characterized by the statistical properties of the probability distributions of step length *λ*(*l*) and turning angle *ψ*(*θ*).

### Simple random walk

The earliest models of movement based on RWs are uncorrelated and unbiased, referred to as simple random walks (SRW). This means that the direction of movement is independent of previous directions moved and completely random [53, 23]. For our modelling purposes, we consider each component of the step vector to be distributed as a zero-centered Gaussian distribution with the same scale parameter *σ*, so that:

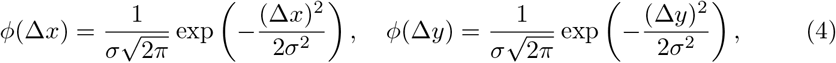

Δ x, Δ ∈ y R, with mean 𝔼 [Δx] = 𝔼 [Δy] = 0 and variance Var[Δx] = Var[Δy] = σ2 which quantifies the mobility of the walker [54].

It can readily be shown that the corresponding step length and turning angle distributions are given by:

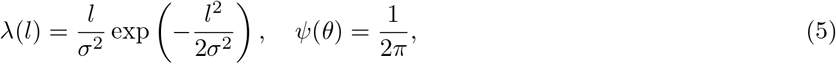

where *λ*(*l*) is the Rayleigh distribution and *ψ*(*θ*) is the uniform distribution ranging from *−π* to *π*, see [52] for a derivation. For this step length distribution, the mean step length and second moment is:

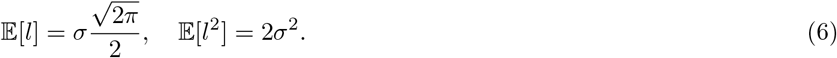

### Correlated random walk

A correlated random walk (CRW) allows for short term directional persistence, so that the movement direction is the same as that of the previous step. For a balanced CRW, the probability of left and right turns are equal, and the turning angle distribution is now considered as a zero centered symmetric circular distribution. An example of such, is the von-Mises distribution:

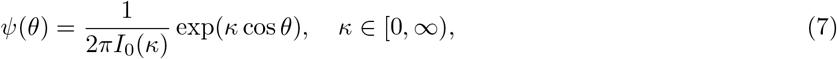

where *κ* is the concentration parameter and *I*_*m*_(*κ*) is the modified Bessel function of the first kind of order *m*, defined through the integral 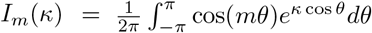. Note that, other types of circular distributions can also be used, for e.g. the wrapped Cauchy or wrapped normal distributions [55]. The mean cosine of the distribution of turning angles quantifies the strength of the short term directional persistence, defined as:

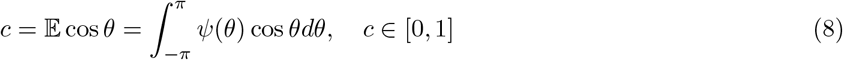

and in the particular case of the von-Mises distribution, this reads:

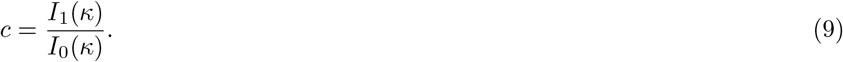

Note that, the SRW corresponds to a special case of the CRW when *c* = 0 (or identically *κ* = 0), as the von-Mises distribution reduces to the uniform distribution, as seen in equation (5). Details of useful metrics which are used to analyse movement patterns, such as the Mean Squared Displacement (MSD) which measures the squared beeline distance between a walkers’ initial and final positions, and the Sinuosity Index (*S*), which measures the amount of turning in a walkers’ movement path, can be found in Appendix.

Figure 1 illustrates a movement path for the SRW and the CRW. With increasing mean cosine *c*, there is an increase in short-term directional persistence, and the path is more diffusive. This demonstrates that the walker is more likely to keep moving in the same direction as that of the previous step, with a decrease in sinuosity.

**Figure 1.**
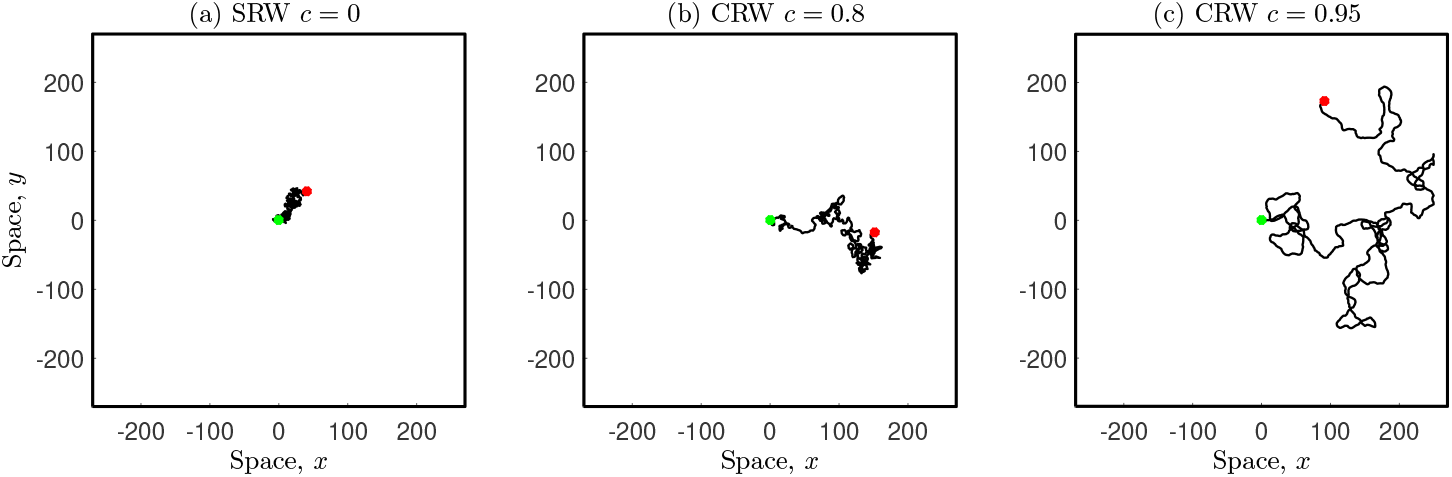
Movement paths for CRWs including the special case of a SRW with *σ* = 1. (a) SRW *c* = 0 (*S* = 1.58) and (b) CRW *c* = 0.8 (*S* = 0.59), (c) CRW *c* = 0.95 (*S* = 0.29). Each walker has starting location at (0, 0) (green marker) and executes *n* = 1000 steps. This corresponds to times *t* = 0 and *t* = 10 using a time increment of Δ*t* = 0.01.

### The correlated Lévy walk

Different types of movement behaviours can be characterized by the rate of asymptotic decay in the end tail of the step length distribution *λ*(*l*). If the end tail decays exponentially or faster (referred to as a ‘thin’ tail), then the variance of step lengths is finite, and therefore the movement process is scale-specific and diffusive [28, 23, 56]. See for example, the case of the SRW with Rayleigh distributed step lengths in equation (5). Lévy walks (LWs) are another conceptual tool used to model movement paths. The main difference between this class of walks, and those prior, is that the end tail decays much more slowly (known as a fat or heavy tail), according to the power law:

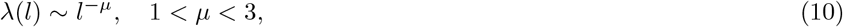

where *µ* is the Lévy exponent. As a result, the walker can execute rare but longer steps, and the movement pattern is much faster. In contrast to RWs with a thin end tail, the variance is divergent, the MSD does not exist and the corresponding movement is scale-free and super-diffusive [57, 38]. Similar to the SRW, the LW is also uncorrelated and unbiased, and the distribution of turn angles is uniform, corresponding to completely random movement. For a correlated Lévy walk (CLW), this distribution is a zero centered circular distribution, where the LW can be considered as a special case.

Without loss of generality, we choose to rely on the folded Cauchy distribution for step lengths:

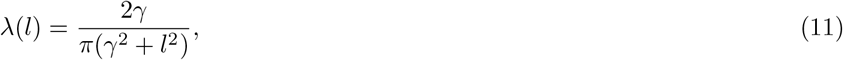

which has quadratic decay in the end tail 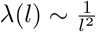 corresponding to Lévy exponent *µ* = 2. Alongside this, we consider the distribution of turn angles to be the von Mises distribution, see equation (7) with mean cosine given by equation (9). The case 0 < *c ≤* 1 now corresponds to a CLW and *c* = 0 to a LW.

To compare between a CRW and CLW, with identical distributions of turn angles, it remains to simply relate *λ*(*l*). This can be done by considering the survival probability ℙ (*l > L*) = *δ*, i.e. the probability of occurrence of move lengths longer than some characteristic scale length *L*, and considering *δ* and *L* to be the same for both distributions. In addition, by imposing an optimization constraint such as minimizing the 𝕃 _2_ norm, one can compute an optimal value for *δ*, and therefore a relationship between scale parameters. As an example, to ‘fairly’ compare between distinct movement types, such as a CRW with step length distribution given by equation (5) and a CLW with distribution given by equation (11), one gets:

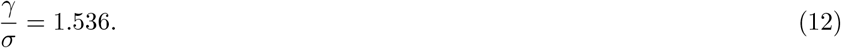

See [58] supplementary information for details of this result.^[1]^

Figure 2 (a) illustrates the movement path of a LW, a specialised type of RW composed of clusters of multiple short steps with longer steps in between them [38]. Plots (b)-(c) show the CLW, which allows for localized directional persistence during the clustering phases, with increase in persistence for larger *c*. In each case, the individual is confined within the square domain, see next section for details of the boundary condition.

**Figure 2.**
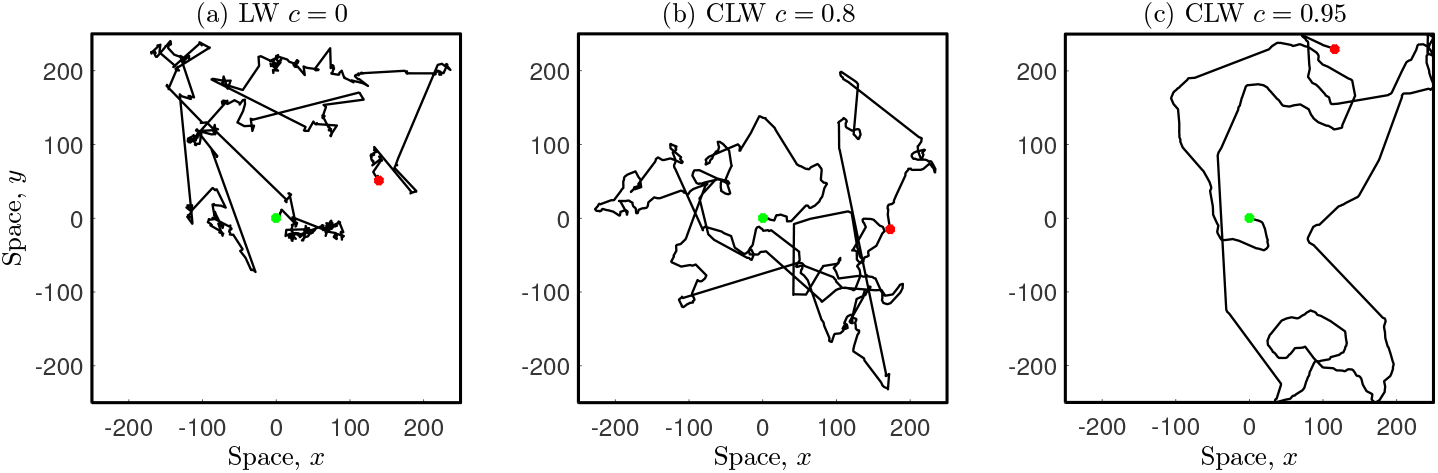
Movement paths for CLWs including the special case of a LW with *γ* = 1.536. (a) LW *c* = 0, (b) CLW *c* = 0.8, (c) CLW *c* = 0.95. Note that, scale parameters can be related through equation (12), so that these RWs are ‘comparable’ to those presented in Figure 1. Each walker has starting location at (0, 0) (green marker) and executes *n* = 1000 steps. This corresponds to times *t* = 0 and *t* = 10 using a time increment of Δ*t* = 0.01

### Simulations

Consider a susceptible population of *N* individuals initially homogeneously distributed across a residential area Ω represented by a square domain of side lengths 2*d*:

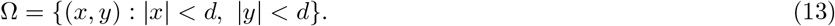

How the population disperses in space can be actualised by modelling the individual movement paths using a *n* step RW, given by the equation:

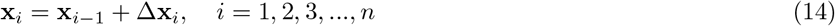

and different movement behaviours can be simulated using the movement rules prescribed by the different types of RWs in *§*. During the course of the movement, individuals may encounter the spatial boundary. We assume that the population is confined, so that no individual(s) can leave or enter, and therefore the domain boundary is considered to be impenetrable. In our simulations, we choose to rely on a ‘no-go’ condition, so that if any individual attempts to overstep the boundary at any instant in time, then an alternative step is chosen at the previous location [59]. In the case that the walk is correlated, a new orientation is assigned in the opposite direction, i.e. in the perpendicular direction to the boundary.

In addition, an infectious host is introduced into the susceptible population at the centre of the domain at location ***ξ***0 = (0, 0), whose movement is modelled as a RW in 2D space:

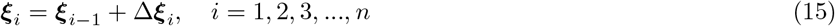

where ***ξ****i* represents the location of the infectious host at each step, and Δ***ξ****i* is a random step vector. Note that, ***ξ****i* and **x**_*i*_ are uncorrelated, so that the movement of the infectious host is completely independent of the movement of any susceptible individual in the population. At each subsequent step, the virus is only transmitted to those susceptible individual(s) that are within a close spatial proximity of less than a distance *r* from the infectious host [21], with condition:

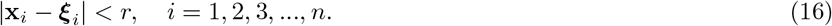

Since the coronavirus is highly infectious, we assume that the probability of disease transmission given a contact is 1. This can be considered as an upper limit of a more general scenario, where host-host contacts can lead to unsuccessful transmission events, known as ‘near misses’. Also, given that our focus is on the short-term dynamics, during the incubation period but prior to the infectious period, we assume that infected individuals do not go on to further transmit the virus. However, the primary infectious host continues to browse throughout the population, as per the RW model, and continues to infect others as a result of further contacts. Levels of infection can be computed as the proportion of individuals that are infected over the course of time.

Figure 3 shows that as the infectious host browses through space, the virus is transmitted to those susceptible individuals in close spatial proximity less than a distance *r* = 10, and therefore the proportion of infected individuals increase over time. The blank space in plots (b)-(d) emerge, as those infected are not shown. Note that, if the residential area is considered to be 500m by 500m, a more realistic value for *r* is 1m, which is used in further simulations, and reflects the current public social-distancing policy in Kuwait.

**Figure 3.**
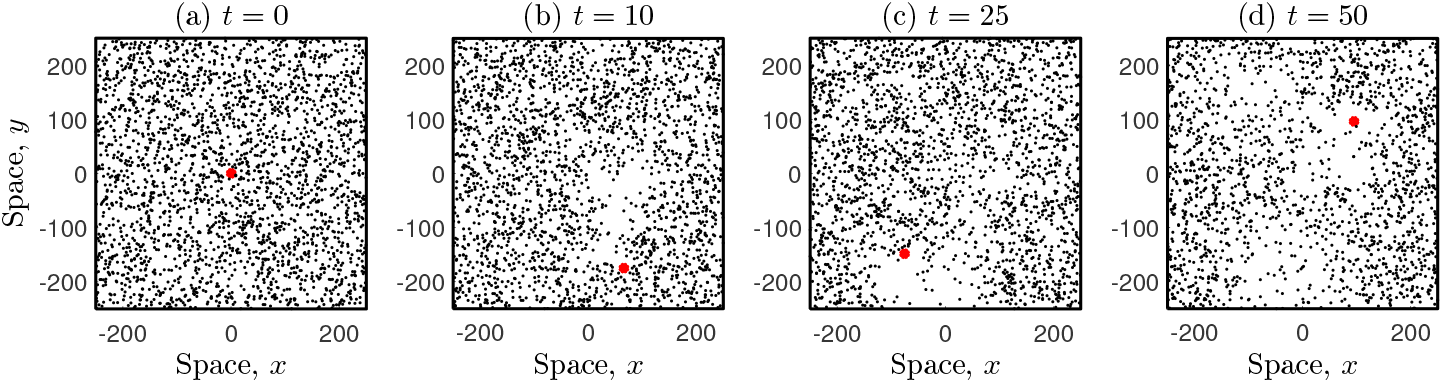
Snapshots of the interplay between an infectious host (red marker) and the population. A susceptible population of *N* = 2500 individuals perform a SRW with *σ* = 1, *c* = 0 within the residential area. The infectious host browses in space according to a CRW with *σ*\* = 1, *c*\* = 0.95, where * is included to distinguish between the parameters. The interaction radius (*r* = 10), movement types and parameters are chosen for illustrative purposes.

## Results

### Variation in infection levels due to host movement

Figure (4) shows that the proportion of infections depend on the interplay between the susceptible population and the infectious host movement. Plot (a) demonstrates that in the case of slow population diffusion (i.e. low values of *c*), infection levels increase at a much faster rate with respect to the directional persistence of the infectious host, but this rate decreases with larger *c*. This suggests a mechanistic explanation for ‘super-spreaders’ based on the movement of the host population as a driver for increasing epidemic spread. In the case of much faster population diffusion (i.e. *c* close to 1), infection levels tend to remain constant - which is also observed if the population is super-diffusive, as seen in Plot (b). Interestingly, the Lévy movement patterns suggest that the rapid movement of individuals cannot explain rising levels of infection.

**Figure 4.**
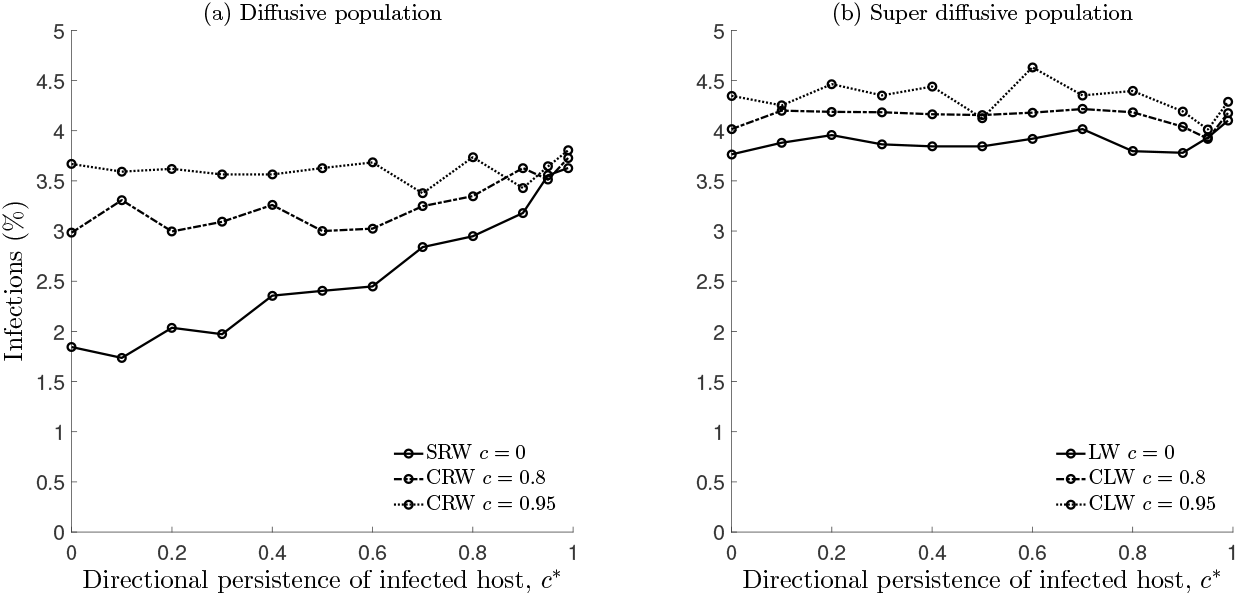
Proportion of infections (%) computed for different scenarios of population dispersal, whilst a single infectious host browses throughout the population, recorded at time *t* = 40. A susceptible population of *N* = 2500 is initially homogeneously distributed across a square domain of side lengths 2*d* = 500, referred to as the residential area. Each individual in the susceptible population moves according to either: (a) SRW or CRW (*σ* = 1), and in (b) LW or CLW (*γ* = 1.536), whilst considering an increase in directional persistence, which is quantified by *c* (indicated in the figure legend). These mobility parameters are related through equation (12)xs, which allows for the proportion of infections to be compared across different host movement types. The infectious host moves according to a CRW with *σ*\* = 1, with varied directional persistence *c*\*. The interaction radius is fixed at *r* = 1. Time is computed as *t* = *n*Δ*t*, with *n* steps in the RW, and time increment Δ*t* = 0.01. Simulations were averaged over ten runs, to reduce the effect of stochastic noise.

### Impact of confined neighbourhoods

Figure 5 (a)-(b) illustrates the free movement of four infectious hosts amongst a homogeneous population. In (c)-(d), confinement restrictions are imposed by separating the residential area into four neighbourhoods, demarcated by solid red lines. The proportion of infections can be computed due to contacts between individuals in the susceptible population and each infectious host. In this manner, we aim to assess the impact of sub-population confinement.

**Figure 5.**
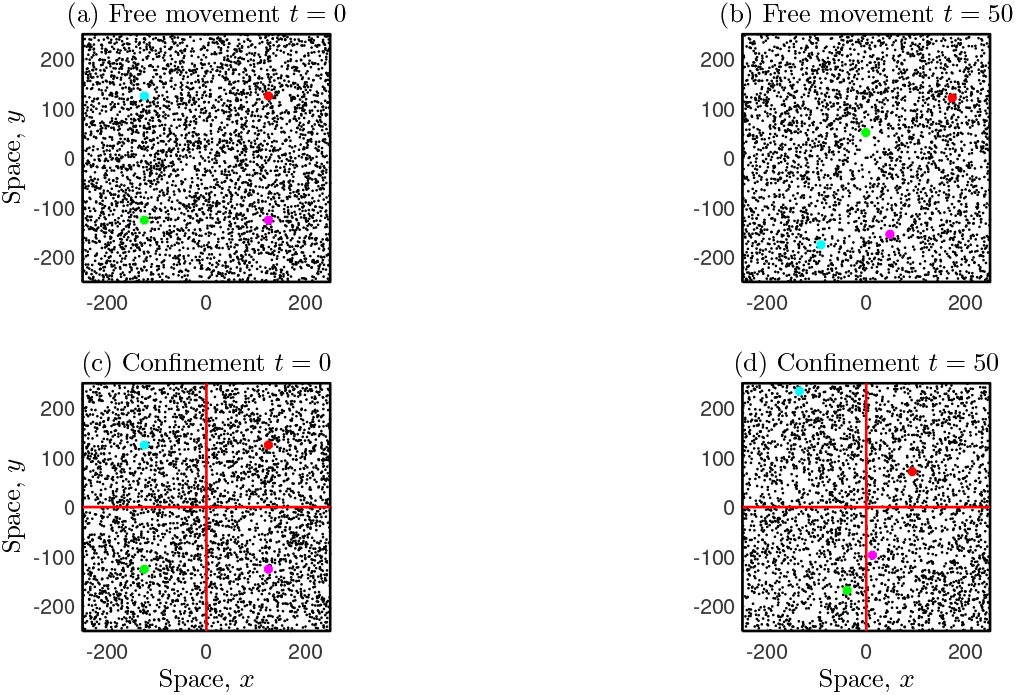
Introduction of neighbourhood confinement. (a) A susceptible population of *N* = 4000 individuals is homogeneously distributed across a residential area with dimensions *d* = 500 by *d* = 500. We consider four infectious individuals at initial locations 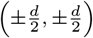. (b) The host population moves freely across the whole domain, where each individual performs a RW. Susceptible individuals that come into close contact of either of these infectious hosts are also infected, with interaction radius *r* = 1. (c)-(d) An alternative scenario is considered where confinement restrictions are imposed, by partitioning the residential area into four neighbourhoods of equal population density. Each sub-population of *N* = 1000 individuals, and also the corresponding infectious host, is thus confined to each neighbourhood of dimensions *d* = 250 by *d* = 250. A ‘no-go’ boundary condition is also prescribed to the inner boundaries, as described in *§*. As a result, potential contacts and hence the transmission of the virus is also confined.

Figure 6 represents a similar scenario, whereas now we consider an overcrowded neighbourhood. Note that in plots (d)-(f), due to the super-diffusive properties of the LW, the sub-population mixes much faster with the remainder of the population, c.f. with plots (a)-(c).

**Figure 6.**
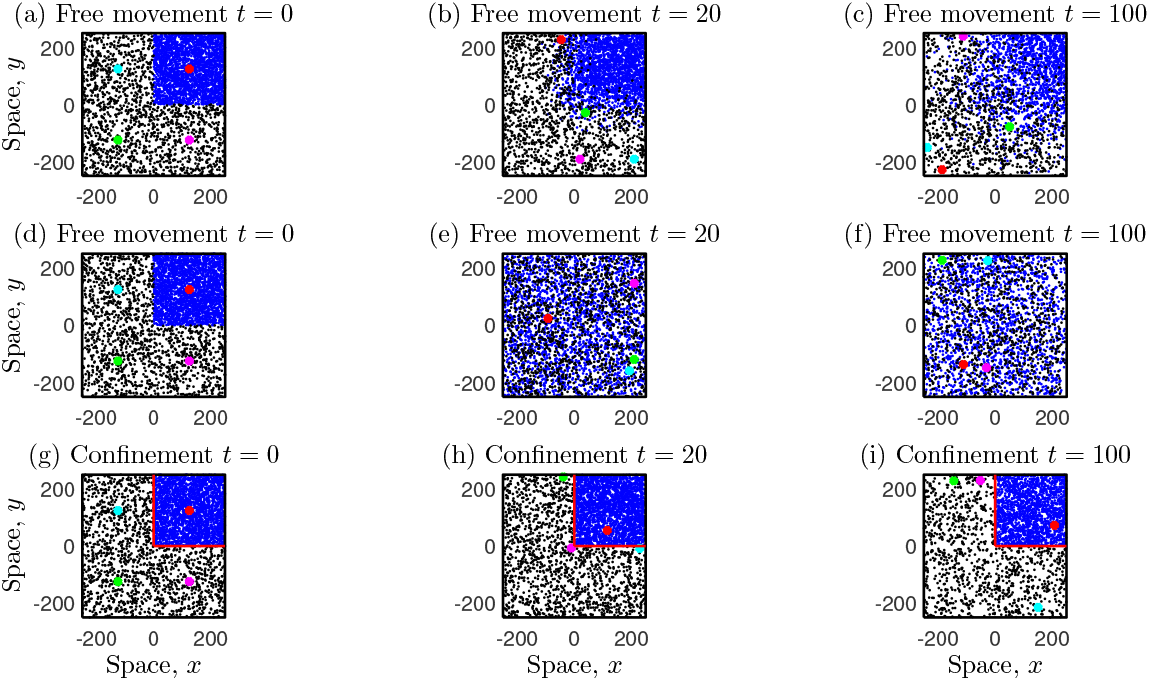
The overcrowded neighbourhood has a population of *N* = 2500 (top-right in each plot indicated in blue), and *N* = 1500 in the remaining part of the residential area, corresponding to an initial high population density which is five-fold. Plots (a)-(c) demonstrate the case where each individual in the susceptible population performs the SRW, and in (d)-(f) the LW. Plots (g)-(i) illustrate confinement of overcrowded area. All other details are the same as that in the caption of Figure 5

Figure 7 plot (a) shows that for susceptible individuals that perform a SRW, the disease spread depends on the movement of the infectious host, with an increase observed for larger *c*\*, however, from plot (c), in the case of a LW population they remain the same - as seen previously in Figure 4. Plots (b) and (d) confirm that this also applies to heterogeneous populations, for e.g. in the case of overcrowded neighbourhoods. This suggests that infection levels depend on the underlying movement behaviours which govern contacts between individuals, but not on the spatial distribution of the host population. On comparing free movement across the whole residential area (solid lines) and restricted movement within confined space (dotted lines), the impact on infection counts is negligible, at least in the short-term.

**Figure 7.**
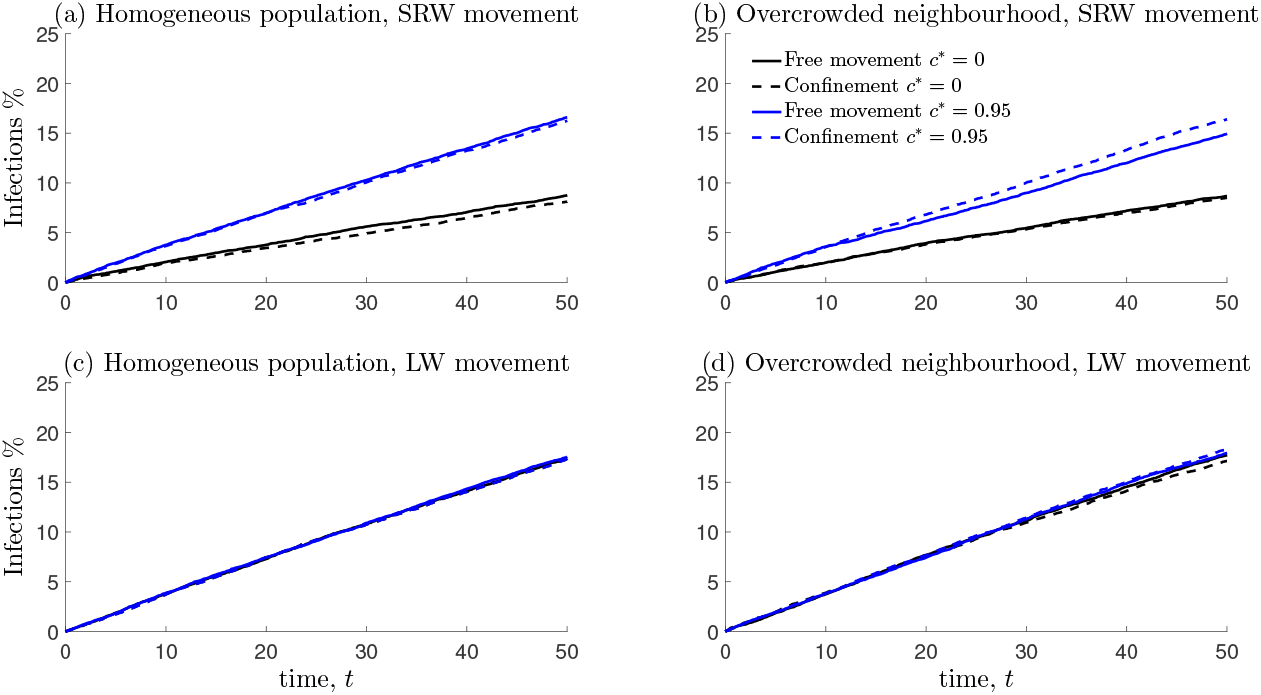
Plot of infection count trajectories against time, whilst considering either a homogeneous population distribution, or the case of an overcrowded sub-population, including the different confinement scenarios depicted in Figures 5 and 6. We also consider two different modes of susceptible population movement, where individuals either perform the SRW or the LW. Each infectious host, browses throughout the population whilst performing a SRW (*c*\* = 0) or a CRW with high directional persistence (*c*\* = 0.95).

## Discussion

Traditional models of infectious diseases usually assume that susceptible and host populations mix readily, and fail to capture the interactions between individuals [60, 61, 62, 63], whereas mechanistic models account for the spatial proximity between individuals and explicitly model the contact process - which is directly related to the disease transmission process [20, 11]. Also, much focus in the literature is on the long term effects of epidemic spread, which is important to address questions related to disease spread/control and the resulting socio-economic impacts etc. [7, 64]. In contrast, very few studies focus on the short time scale of infection dynamics following an outbreak. There is growing attention towards asymptomatic yet infectious carriers, as they are hard to track and could be a critical factor in the spread of some diseases [65, 66, 67], however, not much information is available at very early stages i.e. prior to when individuals become infectious. In this paper, we set out to investigate how infection levels are driven by the interplay between a susceptible population and a single infectious host on a short time scale, whilst considering different types of movement behaviours, heterogeneous population spatial distributions and the role of neighbourhood confinement. An individual mechanistic modelling approach was used based on RWs, which is increasingly recognised as a fundamental tool of infectious disease epidemiology [20, 11].

We found that infection levels increased more rapidly for a slowly diffusive susceptible population with increase in short term persistence in host movement, but remained stable in case of faster or super-diffusive population diffusion. The former may be more relevant in the context of disease ecology with infectious disease spread amongst animals [68, 69, 70, 71], whereas the latter is more applicable to human movement, due to the plausible evidence that human mobility patterns contain statistically similar features observed in super diffusive spread or Lévy walks [45, 46, 47, 48, 49, 72]. This suggests that the rapid movement of humans fails to explain rising levels of infections. We also investigated whether there were any changes in infection levels based on the population spatial distribution (i.e. overcrowded scenarios) and/or neighbourhood confinement. We found that, on a short time scale, there was no noticeable difference.

The state of Kuwait is a typical example amongst many other countries that have implemented strict intervention policies to curb the spread of the disease. The first case was recorded on 24^th^ February 2020. As of July 22^nd^ 2020, the total number of infected cases is 60,434, with 412 deaths, 50,919 recoveries and the number of active cases is currently 9,515 [3, 73]. Some standard approaches such as discontinuation of commercial flights, school closures, social distancing policies, quarantine etc. are common, whilst others are unique to Kuwait, for e.g. extreme household curfew timings up to 22 hours, imposed movement restrictions so that residents can only purchase groceries in their locality subject to prior appointment, total confinement of overcrowded neighbourhoods etc. For the latter, barricades were set up at all entry and exit points and monitored at all times by security officials, ensuring that people were not allowed to leave or enter these areas, except under very special circumstances. See [74] for a detailed timeline of all government interventions. Although, these strict measures have far-reaching consequences beyond the spread of the disease, with clear social-economic impacts [64], the swift response by Kuwait has been recognized and generally praised by the WHO [75]. Given that this strategy is unique, some theoretical questions need to be addressed regarding its efficacy c.f. [76], and whether they can be implemented at larger spatial scales, i.e. it may be challenging or even non-feasible for countries with large populations, particularly in case of cities which are densely populated.

Our results indicate that during the early stages of infectious spread, imposed confinement restrictions or complete lock down of whole residential areas may be ineffective, irrespective of the population demographics, and therefore other types of measures are more likely to be beneficial. On a much larger time scale, lockdowns are known to be effective and there are recommendations that they should remain in place for a time period of about 60 days [77]. This highlights that there is an optimal time frame, precisely when and how a lockdown should be enforced [78, 79]. It is plausible that the exact timing may depend on the population characteristics (movement, spatial structure etc.) and the rate of initial spread, and this would constitute an important research line of enquiry in a future study. Such information is vital for stakeholders (government, health officials, policy makers etc.), as some countries are past their (first) epidemic peak, and a second wave of the pandemic is predicted [80, 81]. One important aspect of this study is that no further disease transmission was assumed once individuals in the susceptible population were infected, which is adequate on a short time scale. However, to investigate the long-term effects, one would need to account for further transmission from newly infected individuals. It would be interesting to analyse the infection dynamics based on population spatial distributions, the interplay between host movements and various confinement scenarios on this time scale. Considering this, one could estimate variations in a key epidemiological metric - the effective reproduction number (*R*), i.e. the average number of secondary cases per infectious case in a population [82]. In terms of contact specifics, we assumed that those individuals who were in close spatial proximity of the infectious host to be instantly infected – which could be justified in the case of highly contagious viruses. A more realistic scenario would assign a transmission probability, so that the virus is transmitted to only a proportion of those individuals who come into close contact. This would allow disease modellers to identify and quantify ‘near misses’ and to explore possible alternative epidemic outcomes given shifts in epidemiological parameters [11]. Moreover, if contact rates and transmission probabilities can be estimated from epidemic/movement data, mechanistic models could prove to provide a powerful modelling framework for a broader category of diseases [11].

## Conclusions

Our findings indicate that infection levels can vary depending on the movement rules that govern host-host movement. In the case of a slowly diffusive susceptible population, the level of infection increases, but remains the same for a highly or super diffusive population. We also found that in the short-term, prior to when susceptible individuals becomes infectious, confinement restrictions are ineffective, even in the case of overcrowded populations. This study demonstrates how useful insights of disease dynamics can be obtained from a random walk framework, in contrast to traditional modelling approaches.

## Data Availability

No data was used.

## Appendix A: Useful metrics for CRWs

The Mean Squared Displacement (MSD), which is defined as the expected value of the squared beeline distance between a walkers’ initial and final positions in a *n* step RW, serves as a useful metric to analyse movement patterns. For a balanced CRW, this can be computed as:

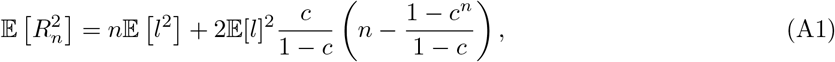

which is expressed in terms of moments of step length *l*, mean cosine *c* and the number of steps *n* [26]. In the special case of a SRW (*c* = 0), the MSD reduces to:

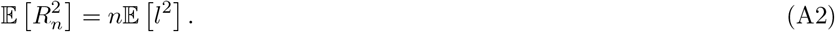

For a large number of steps *n* (or equivalently in the long term), the MSD approaches:

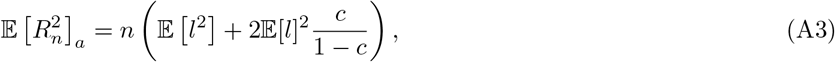

where the subscript ‘*a*’ is included here, to distinguish between the asymptotic MSD and the actual MSD in equation (A1). Note that, the asymptotic MSD grows linearly with *n* and therefore the RW becomes diffusive in the large step limit, and can be related to the diffusion coefficient *D* [83, 84, 85, 23, 86], through the relation:

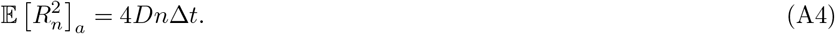

Another useful metric is the sinuosity index *S*, which quantifies the amount of turning in a walkers’ movement path (tortuosity), defined as:

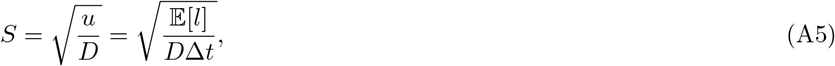

where 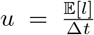 is the mean speed [87]. On combining equations (A4)-(A5), an equivalent expression for the sinuosity index can be written as:

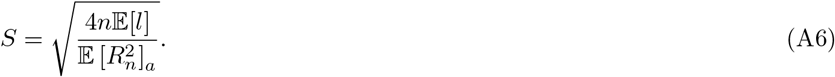

In the particular case of a balanced CRW with Gaussian increments, this index is given by:

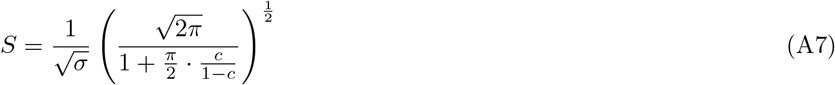

where the moments are computed in equation (6).

## List of abbreviations

WHO: World Health Organization
RW: Random Walk
SRW: Simple Random Walk
CRW: Correlated Random Walk
LW: Lévy Walk
CLW: Correlated Lévy Walk

## Acknowledgements

None

## Authors’ contributions

DAA, ARA, MI and NA conceived the study. DAA conducted the simulations. DAA and KD provided a formal analysis. DA, AA, MI and NA were involved in the funding acquisition. All authors wrote the draft and critically revised the manuscript, and all authors approved the submission.

## Funding

This research was funded by the Kuwait Foundation for the Advancement of Sciences (KFAS) grant number: CORONA PROP 31.

## Availability of data and materials

No data was used.

## Ethics approval and consent to participate

Not applicable.

## Consent for publication

Not applicable.

## Competing interests

All authors have read and agreed to the final version of the manuscript, and declare no conflict of interest.

## Footnotes

[1] [58] compute a relationship between distribution scale-parameters, but consider the probability *ϵ* that move lengths do not exceed *L*, i.e. P(*l* < *L*) = *ϵ* = 1 − *δ*. In either case, the result in equation (12) is the same.

## Notes

### Competing Interest Statement

The authors have declared no competing interest.

